# Host microRNAs are differentially expressed in EBV+ Post-transplant Lymphoproliferative Disorder solid-organ transplant recipients

**DOI:** 10.1101/2022.06.20.22276573

**Authors:** Ayantika Sen, Jeanna Enriquez, Mahil Rao, Marla Glass, Yarl Balachandran, Sharjeel Syed, Clare J. Twist, Kenneth Weinberg, Scott D. Boyd, Daniel Bernstein, Amber Trickey, Dita Gratzinger, Brent Tan, Mary Gay Lapasaran, Mark A. Robien, Merideth Brown, Brian Armstrong, Dev Desai, George Mazariegos, Clifford Chin, Thomas Fishbein, Robert S. Venick, Akin Tekin, Heiner Zimmermann, Ralf U. Trappe, Ioannis Anagnostopoulos, Carlos.O. Esquivel, Olivia M. Martinez, Sheri M. Krams

## Abstract

Post-transplant lymphoproliferative disorder (PTLD) is a serious complication of solid organ transplantation (SOT). Predisposing factors include primary Epstein-Barr virus (EBV) infection, reactivation of EBV in recipient B cells, and decreased T cell immunity due to immunosuppression. Previously, we demonstrated that EBV infection markedly reshapes the microRNA (miR) landscape in EBV+ B cell lines leading to increased IL-10 production. To establish the miRNAome of PTLD tumors we analyzed formalin-fixed, paraffin-embedded shavings of tumor tissues obtained from EBV+ PTLD SOT recipients by microarray analysis and quantitative PCR. The miRNAome of EBV+ PTLD tumors were distinctly different from EBV-PTLD tumors with reduced expression of miRs-17, 19 and 106a, and 194 among EBV+ PTLD tumors. miRs-17, 19, 106a, 155, and 194 were quantitated in the plasma and extracellular vesicles (EVs) from EBV+ PTLD+ SOT recipients and matched transplant controls. The plasma and EV levels of miRs-17, 19, 106a and 194 trended lower in the EBV+ PTLD+ group compared to matched controls, with miR-17 (plasma), miR-19 (EVs) and 106a (plasma and EVs) being significantly reduced. Importantly, the cell free miRs were contained primarily within the EVs. Further studies on the diagnostic, mechanistic, and therapeutic potential of these miRs in PTLD are warranted.

## 1. Introduction

Post-transplant lymphoproliferative disorder (PTLD) is a major complication of immunosuppressive therapy after solid organ transplantation (SOT) and is associated with significant mortality. PTLD includes a heterogeneous group of lymphocytic proliferations, however, a majority of PTLD cases are of B-cell origin and are associated with Epstein-Barr virus (EBV) infection.^1,2^ While there is a clear association between EBV and the development of PTLD, our understanding of how EBV infection leads to PTLD remains incomplete.

EBV is a B-cell lymphotropic γ-herpesvirus that infects more than 90% of the adult population worldwide.^3^ In healthy children, primary infection may either go undetected or be manifested as a mild upper respiratory tract infection. If primary infection occurs in young adults, the symptoms may be more severe, and result in infectious mononucleosis (IM).^4^ During primary infection the virus targets the epithelial cells of the nasopharyngeal region, where it rapidly replicates, lysing the host cells and releasing virions that infect neighboring cells. As the virus reaches the secondary lymphoid tissues, it infects B cells that enter the circulation, releasing more virions which go onto infect other subsets of B cells, including memory B cells. The latent phase is a state of dormancy, where the viral genome exists as a nuclear episome that replicates only once per host cell division with only a distinct subset of viral genes expressed.

The ability of EBV to evade immune surveillance has significant effects in immunocompromised hosts, e.g., SOT recipients. One mechanism that has been identified in immunocompromised hosts is through modulation of host miR expression. miRs are small, non-coding, and highly conserved RNAs that regulate gene expression, cell cycle, apoptosis, immune cell development and differentiation by binding to target mRNA and inhibiting translation of the mRNA.^5,6^ LMP1 and LMP2A upregulate host miR-155 in EBV-associated nasopharyngeal carcinoma (NPC) cell lines and clinical samples.^7^ We have also reported the upregulation of miR-155 and its target FOXO3a by overexpression of LMP1 in B-cell lymphoma cell lines.^8^ Tumorigenic roles of miRs encoded by cell-cycle regulatory gene cluster, miR-17∼92, have been reported. Overexpression of miR-18a, encoded by miR-17∼92, causes tumor formation in EBV-associated NPC cell lines and mouse models of NPC.^9^ Other studies have reported the upregulation of miR-21 (encoded by miR-17∼92), by LMP1 in NPC cell lines^10^ and in EBV-negative diffuse large B cell lymphoma (DLBCL) cell lines transfected with EBNA2.^11^ We previously demonstrated that EBV specifically suppresses miR-194. levels which promotes proliferation of EBV+ B-lymphoma cell lines via increased production of IL-10.^12^ Herein the host miRNAome in PTLD tissues and plasma of EBV+ PTLD patients is analyzed.

## 2. Materials and Methods

### 2.1 Patient samples

#### PTLD tumor sections

Tissue sections were obtained from 24 adult SOT cases of PTLD, diagnosed at the Department of Pathology, Charité Universitätsmedizin Berlin, between June 1994 and May 2013. Corresponding clinical data were obtained from the German PTLD registry. The German PTLD registry is a prospective registry that had been initiated in Germany in 2006 to assess the clinical features, treatment options and outcome of PTLD. Data is collected in before, during and at least at 4 weeks, 6, 12 and 24 months after treatment. The ethics committee of the University Medical Center Schleswig-Holstein, Campus Kiel, Germany approved the registry, and all patients gave written informed consent according to the Declaration of Helsinki. Disease stage at enrollment was determined through a full patient history, physical examination, laboratory investigations (including complete blood count, serum lactate dehydrogenase activity, and renal and liver function tests), bone marrow biopsy, and computed tomography scans of the head, chest, and abdomen. The diagnosis of PTLD was based on the examination of formalin-fixed, paraffin-embedded (FFPE) tissue specimens, obtained either by open biopsy or core needle biopsy. All diagnostic tissue samples including conventional histology (i.e., hematoxylin and eosin and Giemsa stains) and immunohistochemistry were reviewed by an expert pathologist and classified according to the criteria of the 2008 WHO classification^13^. EBV was confirmed in 14 PTLD cases by in-situ hybridization for EBERs **(Table 1)**.

#### Blood samples

Blood was obtained from pediatric SOT recipients enrolled at seven sites in the NIAID-sponsored Clinical Trials of Organ Transplantation in Children (CTOTC)-06 (ClinicalTrials.gov Identifier: NCT02182986), a prospective multi-institutional study intended to identify viral and immune biomarkers of EBV-associated PTLD. This study was approved by the Institutional Review Board at each of these sites: Lucile Packard Children’s Hospital Stanford, CA; University of Texas Southwestern Medical Center, Dallas, TX; University of Pittsburgh Medical Center Children’s Hospital, Pittsburgh, PA; University of California, Los Angeles, CA; University of Miami Health System, Miami, FL; Medstar Georgetown Transplant Institute, Washington, DC; Cincinnati Children’s Hospital Medical Center, University of Cincinnati, Cincinnati, OH. All participants provided written informed consent prior to inclusion in the study. Prospective blood samples were collected (n=4753) at enrollment or transplant, every three months during the first two years, and twice yearly thereafter at the time of primary EBV infection, at PTLD diagnosis and during the intense monitoring post-PTLD diagnosis. Participants were followed for a minimum of 12 months and a maximum of four years based on the time of enrollment. Participants enrolled post-transplant (61%) were enrolled within three years of transplantation. EBV serology was performed at the time of transplantation to determine donor and recipient serostatus. EBV PCR to assess viral load was performed by each center’s local laboratory. The primary endpoint was the development of EBV+ PTLD during the study period with 34 participants reaching the endpoint. Matched controls (1 case:2 controls) were selected from the participant pool according to the following criteria: organ type, confirmed EBV positive status, and post-transplant sample availability proximal to the time post-transplant of PTLD diagnosis for the case. Additionally, for case and control participants who were EBV negative at transplant, controls were identified based on (i) the similarity of post-transplant time-to-first detected EBV DNA (DNAemia) to the corresponding PTLD case and (ii) control sample availability at a similar time post-transplant as the corresponding date of PTLD diagnosis. In this study, based on sample availability and timing, plasma samples obtained from 22 pediatric SOT recipients with biopsy-proven EBV+PTLD and 43 matched-controls were analyzed. The ages of participants ranged between 6 months and 19 years with the average age being 7.1 years. About 55% of the patients were females and 45% were males. The types of organ transplant received by participants were heart (n=17), kidney (n=14), liver (n=22), liver and small intestines (n=2), small intestines (n=3) and total visceral transplant (n=7). Among the PTLD+ patients, nine patients had early lesions and 13 had monomorphic (DLBCL)/polymorphic lesions. The average time of PTLD diagnosis post-transplant was 22.5 months **(Table 2)**. Researchers conducting this study were blinded to the PTLD diagnosis. Plasma was isolated, aliquoted and frozen until analyzed.

### 2.2 RNA Isolation, miR microarray, and qPCR from tissue

Total RNA was isolated from PTLD tissue samples obtained from 24 adult cases of PTLD using the RecoverAll™ Total Nucleic Acid Isolation Kit (Ambion, Austin, TX). For each patient sample, three 10 µm sections of the FFPE tissues were processed according to the manufacturer’s instructions. RNA concentration and purity were determined with the Nanodrop® ND-1000 UV-Vis Spectrophotometer (Thermo Fisher Scientific Inc., Wilmington, DE).

Expression of human miRs was analyzed in 150 ng of total RNA from nine patient samples, categorized as EBV+ (n = 4) or EBV-(n = 5) PTLD by hybridization on Affymetrix’s GeneChip miR Array 4.0 (Stanford Functional Genomics Facility, Stanford, CA). The Bioconductor ‘oligo’ package was used to perform array background subtraction, quantile normalization, and summarization by median polish.^14^ The normalized gene expression dataset was annotated with the ‘pd.mirna.4.0’ annotation library package in R (R Core Team).^14^ The expression data was fit to a linear model using the ‘stats’ package in R (R Core Team). Moderated t-statistics and log-odds of differential expression were calculated using the empirical Bayes method. False discovery rate (FDR) tests were performed with the Benjamini-Hochberg procedure for multiple testing correction in R (R Core Team).

Expression levels of five miRs (miR-17, miR-19, miR-106a, miR-155, and miR-194) were quantitated from the remaining 15 PTLD patient samples (10 EBV+ and 5 EBV-). For each sample, 7.0 ng of purified total RNA was used to synthesize miR-specific cDNA using TaqMan™ MicroRNA Reverse Transcription Kit (4366596, Invitrogen, Waltham, MA) and TaqMan® primers (Thermofisher, Waltham, MA) following manufacturer’s protocol.

Expression of miR-17, miR-19, miR-106a, miR-155 and miR-194 in the patient samples were determined by qPCR. The qPCR reaction mix in each well of a 384-well plate consisted of 0.50 µl of 20X qPCR TaqMan® Primers (Catalog no. 4427975, Thermofisher Scientific, Waltham, MA), hsa-miR-17 (Assay ID: 002308), hsa-miR-19 (Assay ID: 000395), hsa-miR-106a (Assay ID: 002169), hsa-miR-155 (Assay ID: 002623), or hsa-miR-194 (Assay ID: 000493); 2X Maxima Probe/ROX qPCR Master Mix (Catalog no. K0231, Thermofisher Scientific, Waltham, MA). A standard curve was generated using Universal miR Reference Kit (Catalog no. 750700, Agilent Technologies, Santa Clara, CA) to quantify each target miR. All experiments were conducted in triplicate. Mann-Whitney test was performed using GraphPad Prism 9 (GraphPad Software, Inc., La Jolla, CA).

### 2.3 RNA isolation from plasma and qPCR

The mirVana™ PARIS™ Kit (AM1556, Invitrogen, Waltham, MA) was used to isolate total RNA from plasma obtained from 65 pediatric SOT recipients. The plasma samples were treated with cell dissociation buffer supplied with the mirVana™ PARIS™ Kit to capture miRs encapsulated in EVs of the plasma. The remaining steps performed followed the manufacturer’s protocol. Removal of genomic DNA was carried out using TURBO DNA-free™ Kit, AM1907, Invitrogen, Waltham, MA) following the manufacturer’s protocol. RNA samples were purified and concentrated using RNA Clean & Concentrator-25 (R1017, Zymo Research, Irvine, CA) following the manufacturer’s protocol. RNA concentration and purity were determined using SpectraMax® i3x (Molecular Devices, San Jose, CA). Concentrations of miRs were determined by qPCR as detailed above. Two-sample t-test was performed using GraphPad Prism 9 (GraphPad Software, Inc., La Jolla, CA).

### 2.4 Isolation of Extracellular Vesicles from plasma samples

Based on sample availability, EVs were isolated from 250 µl aliquots of plasma collected from 18 EBV+ PTLD+ and 37 EBV+ PTLD-pediatric SOT recipients. Isolation was carried out using ExoQuick Plasma Prep with Thrombin (EXOQ5TM-1 System Biosciences, Palo Alto, CA) following the manufacturer’s protocol. The EV pellets were used for further analysis of miR expression. Supernatants from some samples were retained to analyze miR levels in EV-depleted plasma. The EV pellets and supernatants from some plasma samples were subject to nanoparticle tracking analysis (NTA) to confirm if the isolation of EVs was successful. NTA was performed by System Biosciences, Palo Alto, CA.

### 2.5 RNA isolation from Extracellular Vesicles and qPCR

Total RNA was isolated from EV using the SeraMir Exosome RNA Purification Column Kit (Catalog no. RA808A-1, System Biosciences, Palo Alto, CA) following the manufacturer’s protocol. Genomic DNA was removed from isolated RNA samples using TURBO DNA-free™ Kit (AM1907, Invitrogen, Waltham, MA) following the manufacturer’s protocol. RNA samples were purified and concentrated using RNA Clean & Concentrator-25 (R1017, Zymo Research, Irvine, CA) following the manufacturer’s protocol. Concentration and purity were determined using SpectraMax® i3x (Molecular Devices, San Jose, CA). Expression of miR-17, miR-19, miR-106a, miR-155 and miR-194 in the patient samples were determined by qPCR as detailed above. Mann-Whitney test was performed using GraphPad Prism 9 (GraphPad Software, Inc., La Jolla, CA).

## 3. Results

### 3.1 Human miRNAs are differentially expressed in EBV+ and EBV-PTLD tumors

Total RNA was isolated from shavings from 24 FFPE tumor blocks, 14 EBV+ and 10 EBV-, from subjects diagnosed with PTLD (**Figure 1A**). Microarray analysis of host miRs from four EBV+PTLD and five EBV-PTLD, demonstrated that EBV+ PTLD has a distinct miR expression profile as compared to EBV-PTLD (**Figure 1B**). Based on our previous studies we analyzed RNA from the remaining 10 EBV+ and 5 EBV-FFPE PTLD tumor blocks for quantitative expression of miRs-17, 19, 106a, 155 and 194 ^8,12^ (**Figure 1C**). MiRs-17, 19, 106a and 194 were lower in EBV+ PTLD tumors, with miR-194 being significantly decreased (p = 0.0007) in EBV+ tumors. There were no differences in the expression of miR-155 between EBV+ and EBV-PTLD tumors. These results suggest that EBV may modulate expression of host miRs in PTLD tumors.

**Figure 1:**
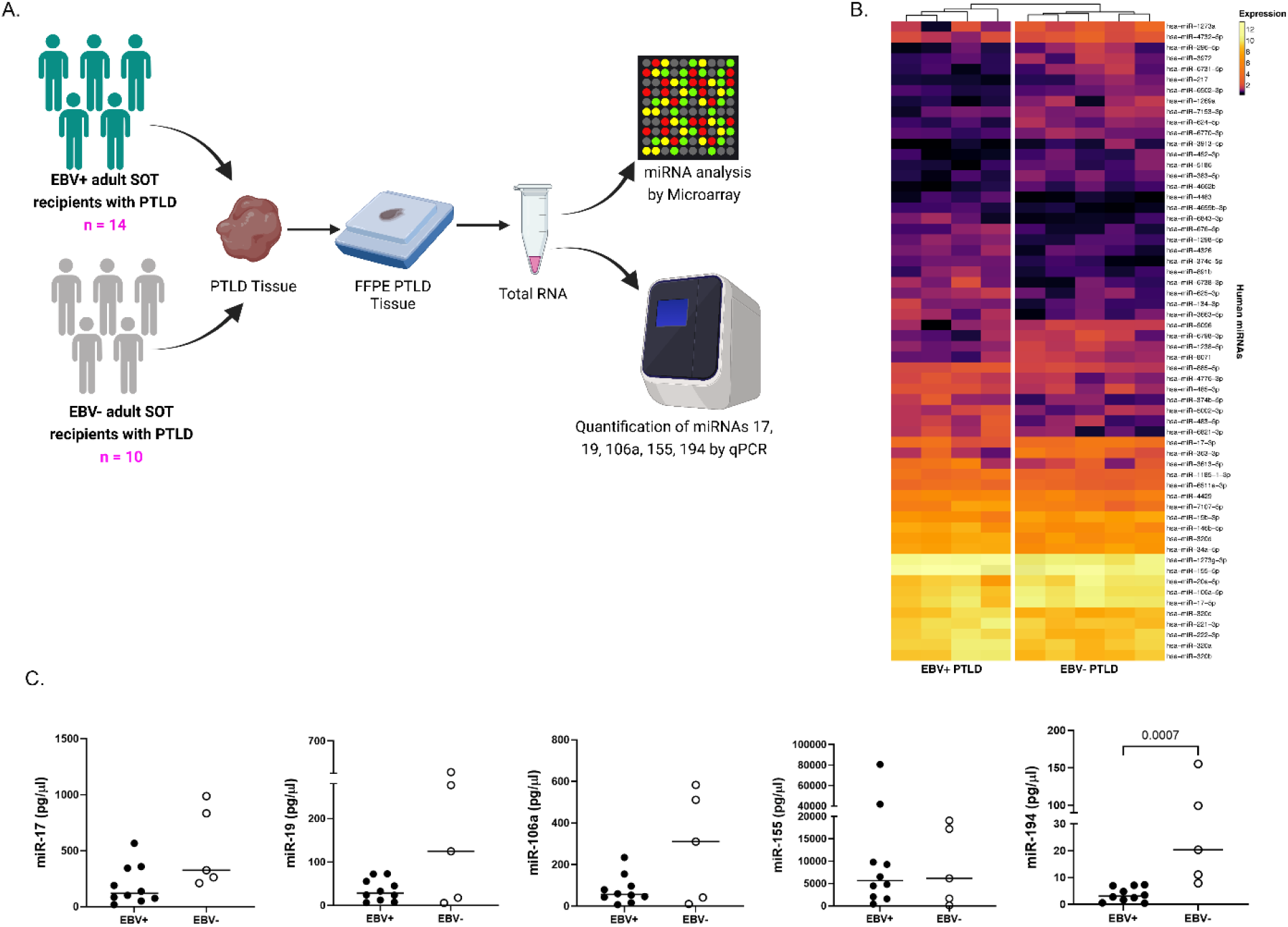
Human miRs are differentially expressed in EBV+ and EBV-PTLD tumors. **(A)** Schematic diagram of the workflow for miR analysis of FFPE PTLD tumors from EBV+ and EBV-solid organ transplant recipient. **(B)** Heatmap showing normalized expression values for a subset of human miRs from EBV+ (n = 4) and EBV-(n = 5) PTLD lesions. **(C)** Concentrations of miR-17, miR-19, miR-106a, miR-155 and miR-194 in PTLD tissues were measured by qPCR in a separate cohort of EBV+ (n = 10) and EBV-(n = 5) patients. miR-194, EBV+ vs. EBV-, p = 0.0007 by Mann-Whitney test.

### 3.2 miR-17 and miR-106a are significantly reduced in the plasma of EBV+ PTLD+ pediatric transplant recipients

We sought to determine if miRs-17, 19, 106a, 155 and 194 were differentially expressed in the circulation of pediatric transplant recipients based on PTLD status, thus we analyzed plasma samples from EBV+ pediatric transplant recipients with and without PTLD. PTLD+ cases (n = 18) were matched 1:2 to PTLD-control (n = 37) pediatric transplant recipients by organ transplanted, EBV serostatus, and the time post-transplant **(Figure 2A)**. The plasma levels of miRs-17, 19, 106a, and 194 were reduced in pediatric transplant recipients with PTLD as compared to matched controls, with miRs-17 (p = 0.034) and 106a (p = 0.007) being significantly reduced **(Figure 2B)**. MiRs-155 expression in plasma did not differ significantly between PTLD+ patients and controls. Further we observed lower expression of miRs-17 and -106 in pediatric transplant recipients with monomorphic/DLBCL or polymorphic lesions as compared to early lesions and controls with miR-106a being significantly reduced (p = 0.047) as compared to matched controls **(Figure S1)**. Pediatric heart and kidney transplant recipients with PTLD demonstrated a marked trend toward lower levels of plasma miRs-17, 19, 106a, and 194 although this did not reach statistical significance since the numbers of samples were lower in each group **(Figure S2)**. Importantly miR-106 was lower in all four groups of pediatric transplant recipients with PTLD, consistent with the significant decrease seen with the combined groups as shown in **Figure 2B** suggesting the importance of this miRNA in PTLD irrespective of organ transplanted. The levels of plasma-derived miRs mirror what we observed in PTLD tumors suggesting the regulated expression of these miRs is in the circulation.

**Figure 2:**
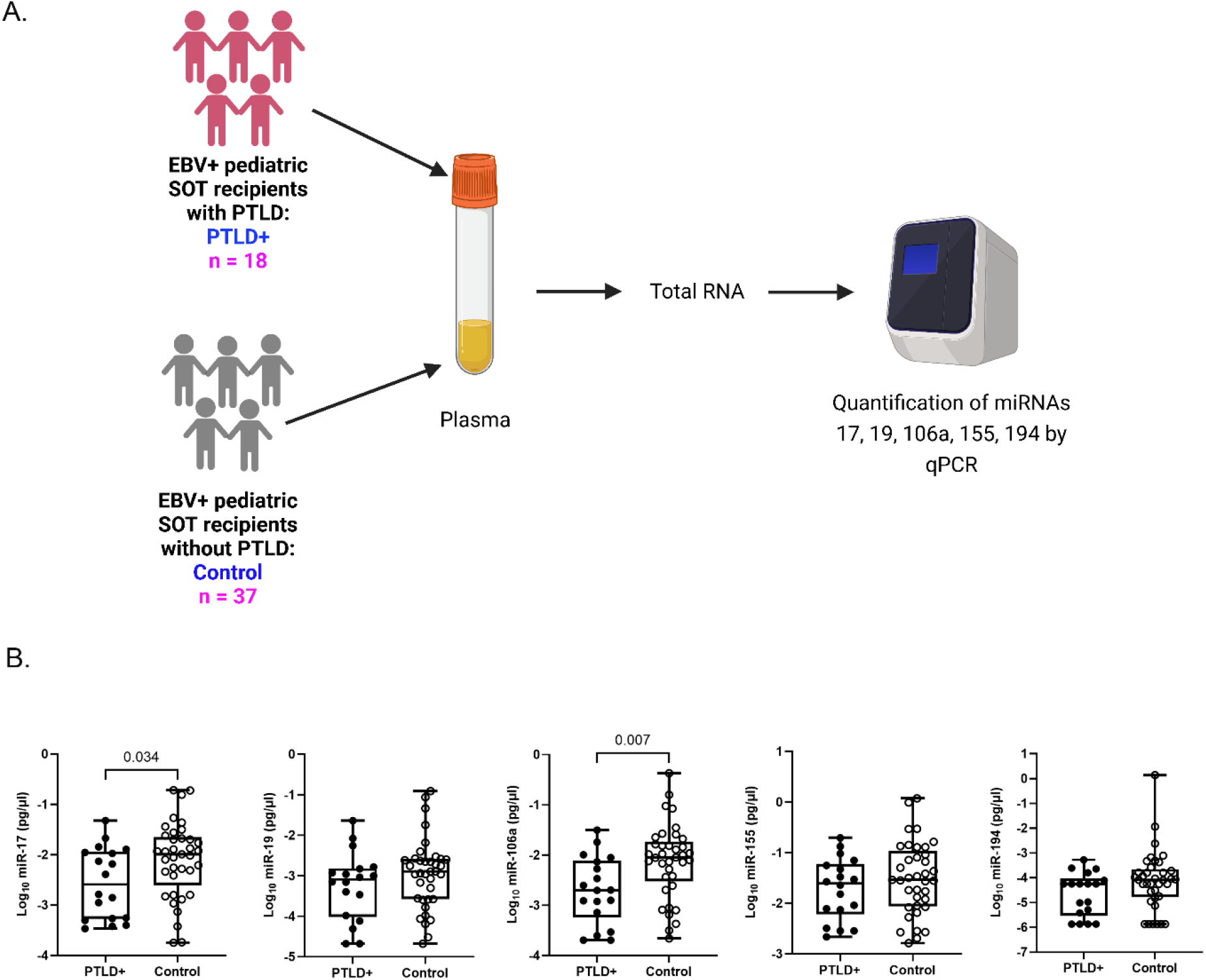
Decreased levels of miRs are detected in the plasma of PTLD+ pediatric transplant recipients as compared to PTLD-pediatric transplant recipients. **(A)** Schematic diagram of the workflow for quantitation of plasma derived miR from PTLD+ pediatric SOT recipients (n = 18) and matched controls (n = 37) **(B)** Concentrations of miR-17, miR-19, miR-106a, miR-155 and miR-194 were measured by qPCR in plasma of PTLD+ and matched controls. miR-17, PTLD+ vs. control, p = 0.034; miR-106a, PTLD+ vs. control, p = 0.007 by two-sample t-test.

### 3.3 Longitudinal analysis of miR expression in plasma indicates a decrease in miR prior to PTLD diagnosis

Longitudinal analysis for miRs-17, 19, 106a, and 194 was performed to determine miRNA expression changes between transplantation and PTLD diagnosis. EBV+ PTLD+ samples (n=13) were matched to controls (n=13) by transplant organ and number of days post-transplant **(Figure 3A)**. From this cohort, five PTLD+ and five control samples having at least three timepoints and the last timepoint being within 60 days prior to PTLD diagnosis (for the PTLD+ group) are shown in **Figures 3B** and **3C**. We observed a decline in plasma levels of miRs-17, miR-19 and miR-194 in all five EBV+ PTLD+ patients at the timepoint closest to PTLD diagnosis. The plasma levels of miR-106a and miR-155 also declined in four out of five PTLD+ patients. Conversely, in four out of five controls the plasma levels of miRs-17, 19 and 106a increased at the timepoint matched with the last timepoint before the PTLD diagnosis in PTLD+ cases.

**Figure 3:**
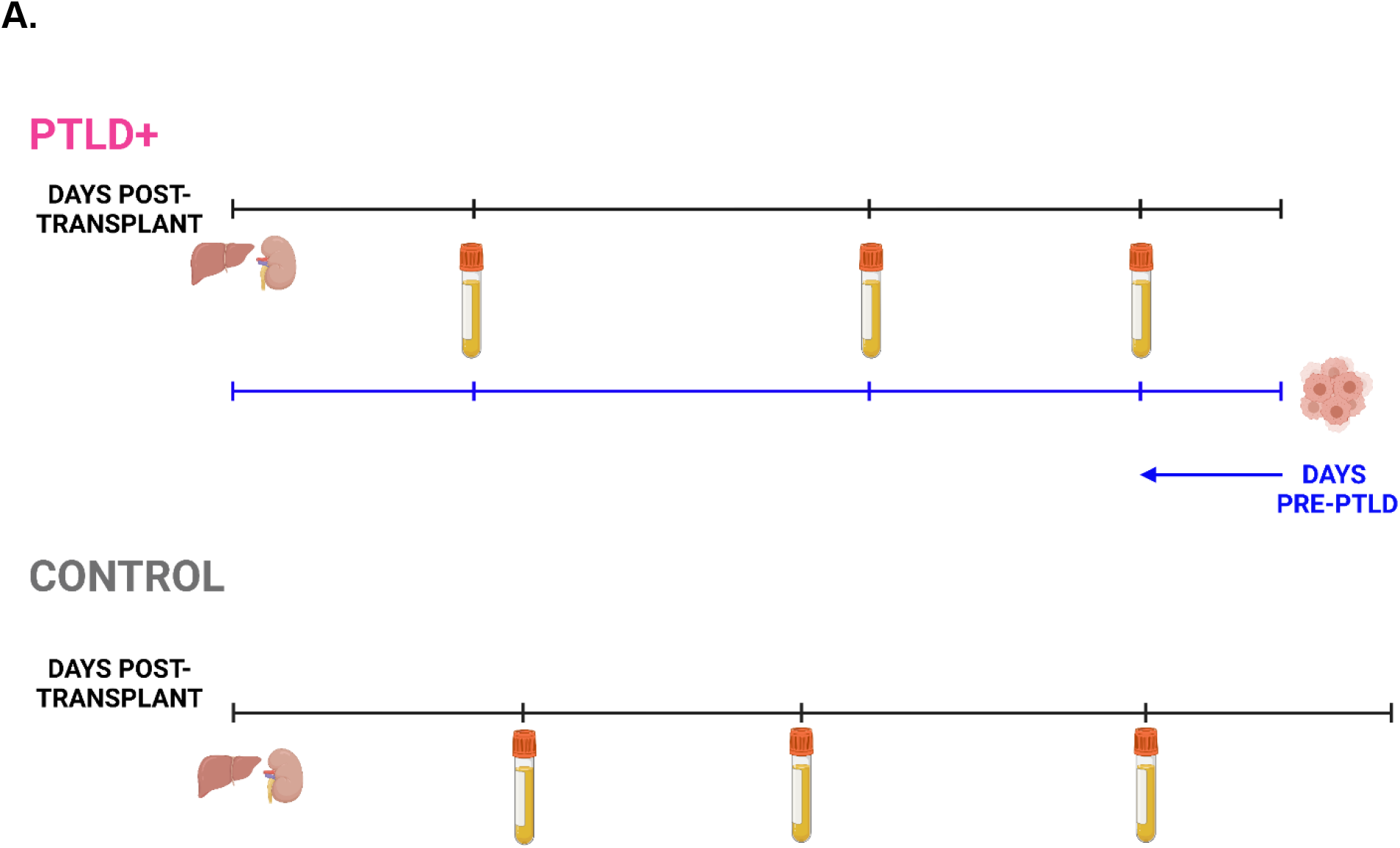
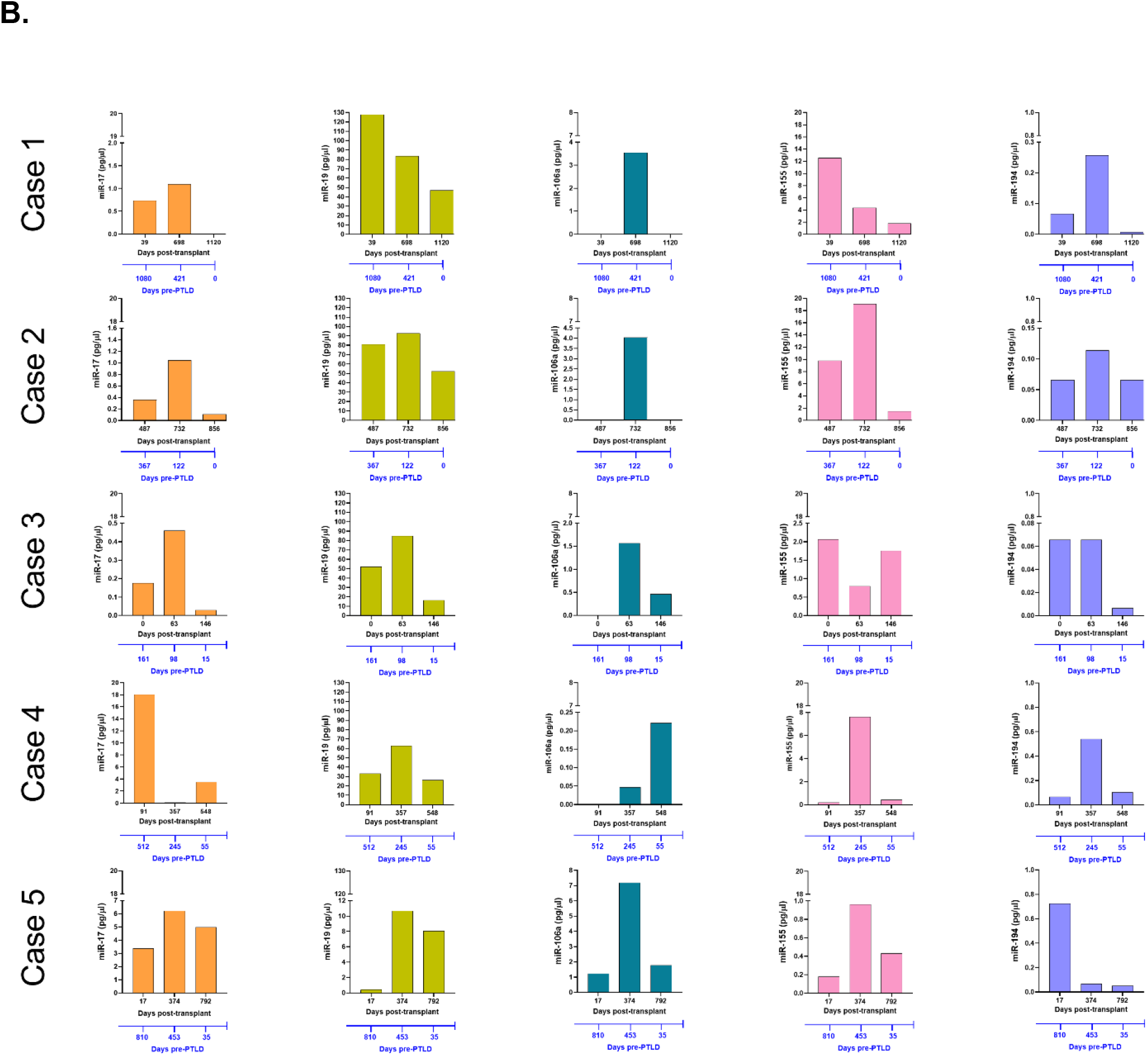
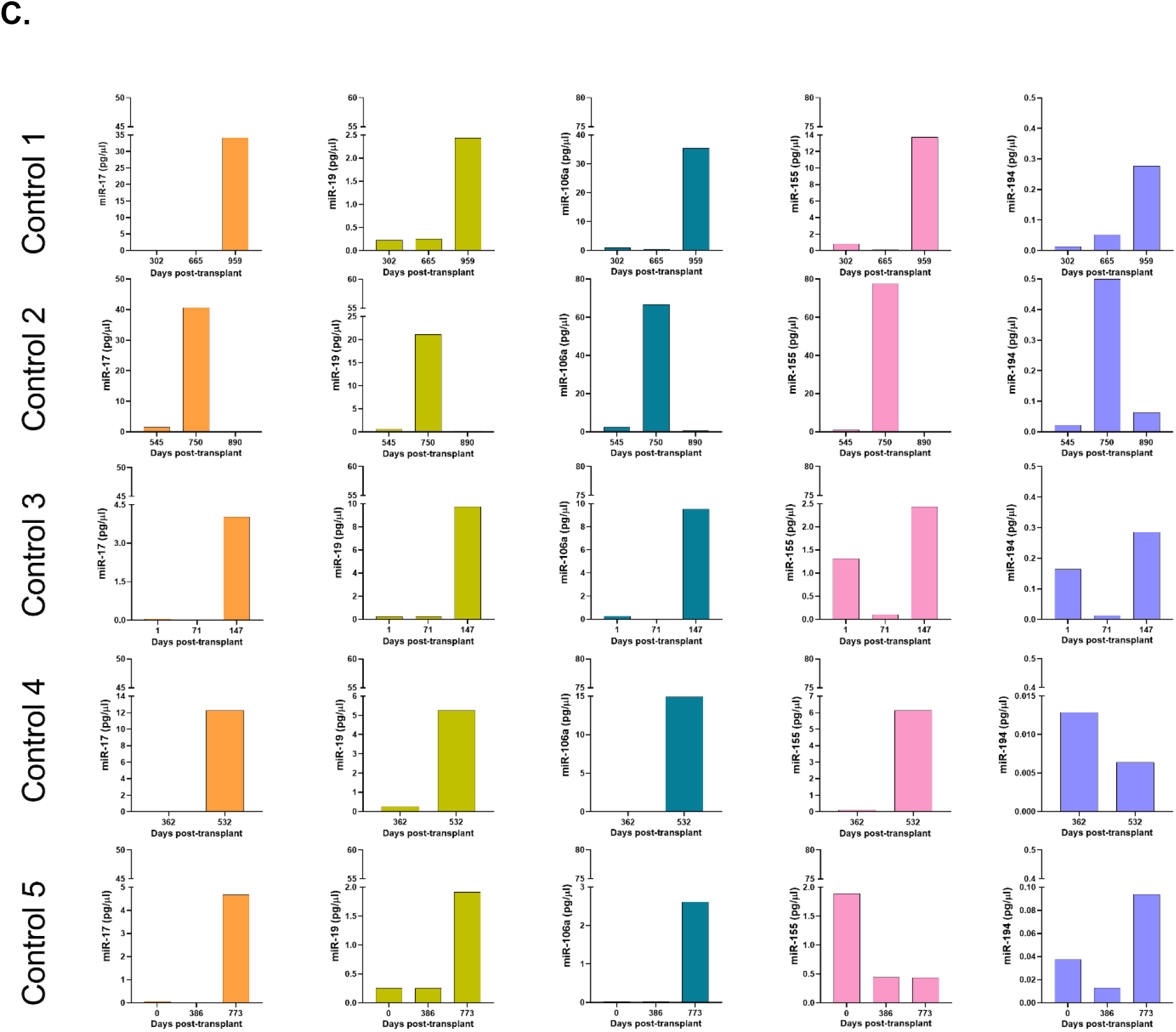
Longitudinal analysis demonstrates a decrease in miR levels prior to a diagnosis of PTLD. **(A)** Schematic diagram of the pairing of PTLD+ (n = 13) and control (n = 13) subjects by matching organ and time of sample collection post-transplant. In both PTLD and control groups, the timeline in black represents the number of days post-transplant. In the PTLD group, the timeline in blue represents the number of days prior to PTLD diagnosis, indicated by Day 0. In the PTLD+ group, the last samples were collected between 0-60 days prior to PTLD diagnosis. **(B)** Concentrations of miR-17, miR-19, miR-106a, miR-155 and miR-194 were measured by qPCR in plasma samples collected at three timepoints post-transplant with the last sample collected within 0-60 days prior to PTLD diagnosis (n = 5). **(C)** Concentrations of miR-17, miR-19, miR-106a, miR-155 and miR-194 were measured by qPCR in plasma samples collected post-transplant from control patients (n = 5) at timepoints matched with paired PTLD+ subjects.

The remaining eight PTLD+ and eight control patients **(Figure S3)** either had samples collected at only two timepoints or had the last sample collected more than 60 days prior to the day of PTLD diagnosis. The data demonstrated a consistent decrease in plasma miR-17 and miR-19 levels and the samples obtained within 60 days of PTLD diagnosis had the least miR-17 and miR-19 levels. These data suggest that miRs 17, 19, 106a and 194 are decreased prior to a clinical diagnosis of PTLD in EBV+ pediatric transplant recipients.

### 3.4 Significantly lower levels of miR-19 and miR-106a are detected in the extracellular vesicles of EBV+ PTLD+ pediatric transplant recipients

miRs produced by cells can be released into the circulation as either free miRs or encapsulated into EVs, which are then released into the circulation. We quantitated the levels of miRs-17, 19, 106a, 155 and 194 in EVs isolated from plasma samples of PTLD+ (n=18) and control patients (n=36) (**Figure 4A**). The mean size of EVs in our samples, as determined by NTA, was 60.1 +/-14.86 nm, consistent with the size distribution of intact exosomes **(Figure 4B)**. The EV-depleted plasma demonstrated a thousand-fold decrease in the concentration of detected particles as compared to the concentration of detected particles in isolated EV pellets.

**Figure 4:**
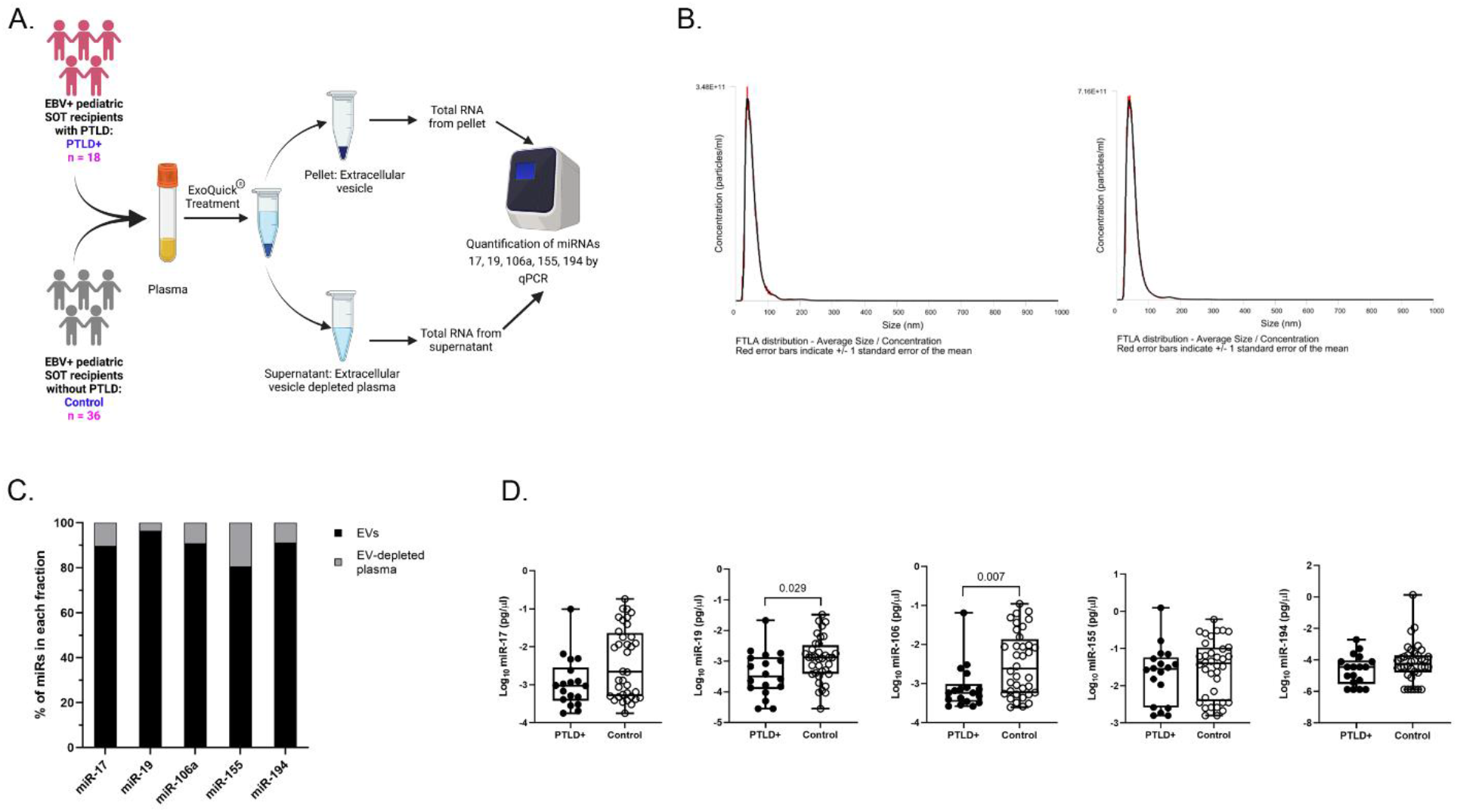
miRs are localized to EVs in plasma. **(A)** Schematic diagram of the workflow for the isolation of EVs from plasma and qPCR analysis of EV-derived miRNA from PTLD (n = 18) and control (n = 36) pediatric SOT recipients **(B)** Characterization of vesicles size and concentration of EVs in EV pellets isolated from plasma fraction by nanoparticle tracking analysis. **(C)** Concentrations of miR-17, miR-19, miR-106a, miR-155 and miR-194 were measured by qPCR in EVs and EV-depleted plasma samples. Black bars represent the fraction of EV-derived miRNAs in whole plasma. Gray bars represent the fraction of miRNAs derived from EV-depleted plasma. **(D)** Concentrations of miR-17, miR-19, miR-106a, miR-155 and miR-194 were measured by qPCR in isolated EVs from PTLD+ (n = 18) and control (n = 36) patients. miR-19, PTLD+ vs. PTLD-, p = 0.029; miR-106a, PTLD+ vs. PTLD, p = 0.007 by Mann-Whitney test.

To determine the proportion of miRNAs sorted into EVs we quantitated the levels of miRs-17, 19, 106a, 155 and 194 in both the EVs and EV-depleted plasma fractions. EVs contained 90% or more of miR-17,-19,-106a, and -194 and 80% of miR-155 **(Figure 4C)**, thus indicating that the majority of the cell-free miRs are sorted into circulating EVs. When we compared the expression of the EV-derived miRs between PTLD+ and control groups, we observed a significant reduction in miRs-19 (p = 0.029) and -106a (p = 0.007) levels in PTLD+ patients while miRs-17 and -194 also trended lower than controls. Expression of miR-155 remained unchanged between PTLD and control groups **(Figure 4D)**. We observed significant decreases in expression of miRs-17 (p = 0.039), -19 (p = 0.044), and -106 (p = 0.009) in the EVs of patients with monomorphic/polymorphic lesions as compared to controls **(Figure S4)**. Taken together, our results indicate that miRs 17,19,106a, and 194, are primarily in circulating EV and are decreased during the development of PTLD in EBV+ pediatric SOT recipients.

## 4. Discussion

We have demonstrated that miR-17, miR-19, miR-106a and miR-194 are decreased in EBV+ PTLD tumors and in circulating EVs of pediatric SOT recipients with PTLD. The data on miR expression in PTLD are very limited. A comparative analysis of miR expression between central nervous system-associated PTLD tissues and systemic DLBCL-like PTLD tissues in adult SOT transplant recipients identified a number of miRs, including miRs-17, 19, and 155, that were differentially expressed between the two cohorts.^15^ To date, however, there has not been an analysis of miR expression in plasma or EVs of SOT recipients with PTLD.

Some forms of PTLD can share histologic characteristics with non-transplant EBV-associated diffuse large B cell lymphoma (DLBCL) or Burkitt’s lymphoma, and there is a growing body of literature on the role of miRs in these histologically-similar lymphomas. miRs-17, 19a, and 106a are encoded by the miR-17∼92 family of gene clusters which includes the miR-17∼92 cluster and its paralogs miR-106a∼363 and miR-106b∼25.^16 17,18^ Expression analysis of human lymphoma samples (both DLBCL and non-DLBCL) revealed upregulation of the miR-17∼92 cluster in the lymphoma tissues.^19^ In another study, 25% of tumor tissues isolated from patients with DLBCL had upregulation of miR-19b compared to patients with reactive lymphoid hyperplasia, while among patients with various types of non-Hodgkin’s lymphoma, higher expression of miR-19b was associated with a reduction in both overall survival and event-free survival.^20^

In our study, however, we observed lower levels of miRs-17 and 19 in patients with PTLD compared to controls. This may be because our patients were receiving immunosuppressants and therefore may had less vigorous T cell immunity to EBV infection. Overexpression of EBV in the AGS gastric carcinoma cell line is associated with reduced expression of the miR-17∼92 cluster via activity of the EBV miR BART1-3p and host transcription factor E2F3.^21^ We too, observed that among patients with PTLD, expression of miRs-17, 19, and -106 were all lower in EBV+ PTLD patients compared to EBV-PTLD patients. Although increased expression of the miR-17∼92 cluster has been associated with aggressive tumor growth, an alternative explanation is miR-17∼92 cluster expression exerts a “Goldilocks”-type effect, with both sub-physiologic and supra-physiologic levels promoting tumor growth.

The sequence for miR-106a is carried by the miR-106a∼363 cluster,^22^ a paralog to the miR-17∼92 cluster, that has been shown to be regulated by common transcription factors and have overlapping functions with miR-17∼92 cluster.^16,23,24^ There are limited studies on the role of miR-106a in PTLD development. However, in a comparative analysis of miR expression between DLBCL tissue, follicular lymphoma (FL) tissue, and healthy lymph nodes, miR-106a expression was significantly higher in DLBCL compared to normal tissue.^25^ The same study also demonstrated that miR-106a was a top driver of differences in expression patterns between healthy lymph tissues and lymphoma tissues. We, however, observed the opposite effect in our samples; miR-106a expression was lower in PTLD+ patients compared to controls. One possible explanation for this is genetic heterogeneity among histologically-similar tumors. Microarray analysis of miR content from DLBCL tumor samples could divide the histologically-similar DLBCL samples into three genetically-distinct groups with differing amounts of miR-106a expression. Samples with lower miR-106a expression also had lower levels of miR-17 and 19, suggesting that the PTLD tissues used in our study may have more similarities to this subset of DLBCL.^26^

DLBCL can be divided into two types based on gene expression profiles, germinal center-like (GC) or activated memory-B cell-like (non-GC), with the GC group having a significantly better prognosis compared to non-GC. miR-155 expression levels were lower in the GC group compared to the non-GC group.^27^ Among children and young adults with Burkitt’s lymphoma, however, the data on miR-155 expression is conflicting. While one study reported high levels of miR-155 in primary Burkitt’s lymphoma tissue, ^28^ others reported that majority of primary Burkitt’s lymphoma tissue did not express miR-155 at detectable levels.^29^ We did not observe any differences in miR-155 expression in EBV+ PTLD tumors or in the plasma. This may also be the result of immunosuppression therapies, that have been reported to suppress miR-155 expression.^30^

The role of miR-194 in EBV-associated cancers and EBV infection is poorly understood. Previously our group reported the EBV-induced regulation of miR-194 in B-cell lymphoma cell lines derived from patients with PTLD.^12^ In this *in vitro* study, miR-194 was found to be downregulated in five out of six PTLD patient-derived EBV transformed B-cell lymphoma cell lines. Analysis by TargetScan and miRanda showed that miR-194, miR-17 and miR-106a have target sites in the 3’UTR region of *il-10* gene and we found that overexpression of miR-194 reduced IL-10 production. Our previous study showed that IL-10 serves as an autocrine growth factor in EBV+ lymphoblastoid cell lines derived from patients with EBV+ PTLD.^31^ We have also shown that inducing LMP1 signaling in EBV-negative B-cells increased the production of IL-10 in these cell lines.^32^ Another study also reported a significantly increased serum IL-10 levels in EBV positive patients at the time of PTLD diagnosis.^33^ Taken together, these studies suggest that EBV can suppress miR-194 expression, leading to increased production of IL-10 and survival of B-cell lymphoma cell lines. Our findings also show reduced expression of miR-194 in PTLD tissues and plasma of EBV+ PTLD patients compared to EBV-PTLD patients.

One of the mechanisms by which miRs can travel through the circulation to distant sites is through encapsulation in EVs. EVs are cell-derived lipid-bilayer nanostructures that carry proteins, lipids and various RNA subtypes from infected cells to remote locations^34^ where they fuse with target cells to transfer their miR cargo. These transferred miRs alter gene expression patterns in the target cells, leading to aberrant growth regulation. EBV is known to exploit EVs to serve as carriers for EBV proteins and miRs. When EVs from EBV+ LCLs were labeled with a fluorescent dye and co-cultured with primary immature monocyte-derived dendritic cells, there was an increase in levels of EBV miRs in the dendritic cells; furthermore, HeLa cells co-cultured with EVs from EBV+ LCLs demonstrated repression of EBV target genes.^35-39^ Our data demonstrate that host miRs can similarly be packaged into EVs and suggest another mechanism by which EBV may regulate gene expression in uninfected cells. These changes in gene expression may have functional consequences for tumor growth as EVs carrying LMP1 from EBV infected DG75 Burkitt’s lymphoma cell lines induce cell proliferation in healthy B cells.^40^

In summary, we demonstrate that selected miRs with potential to regulate growth patterns are differentially expressed in both tumor tissues and plasma of PTLD+ patients. We also demonstrate that these miRs are found in EVs and can travel to distant sites. Lastly, we demonstrate that decreases in expression of miRs-17, 19, 106a and 194 are temporarily associated with the development of PTLD. We have only examined a small number of host miRs which may be altered in the context of PTLD development. Future studies should focus on a thorough characterization of the contents of the EVs in patients with PTLD to determine which host miRs are critical for PTLD development, as miRs may serve as biomarkers for development of PTLD as well as targets for novel therapies.

## Data Availability

The data for this study will be available from the corresponding author upon reasonable request.

## Abbreviations

miR: microRNA
EBV: Epstein-Barr Virus
PTLD: Post-Transplant Lymphoproliferative Disorder
SOT: Solid Organ Transplant
EV: Extracellular Vesicles
qPCR: Quantitative Polymerase Chain Reaction
CTOTC: Clinical Trials of Organ Transplantation in Children
FFPE: Formalin-Fixed, Paraffin-Embedded
DLBCL: Diffuse large B cell lymphoma

## Acknowledgements/Funding

This study was funded by NIH A1UO1AI104342 and UM2AI117870 (Rho).

## Data availability Statement

The data for this study will be available from the corresponding author upon reasonable request.

