## Supplementary material for "Host microRNAs are differentially expressed in EBV+ Post-transplant Lymphoproliferative Disorder solid-organ transplant recipients": Figure S1, Figure S2, Figure S3, Figure S4

Supplemental Document:

**<u>Figure S1:</u>**

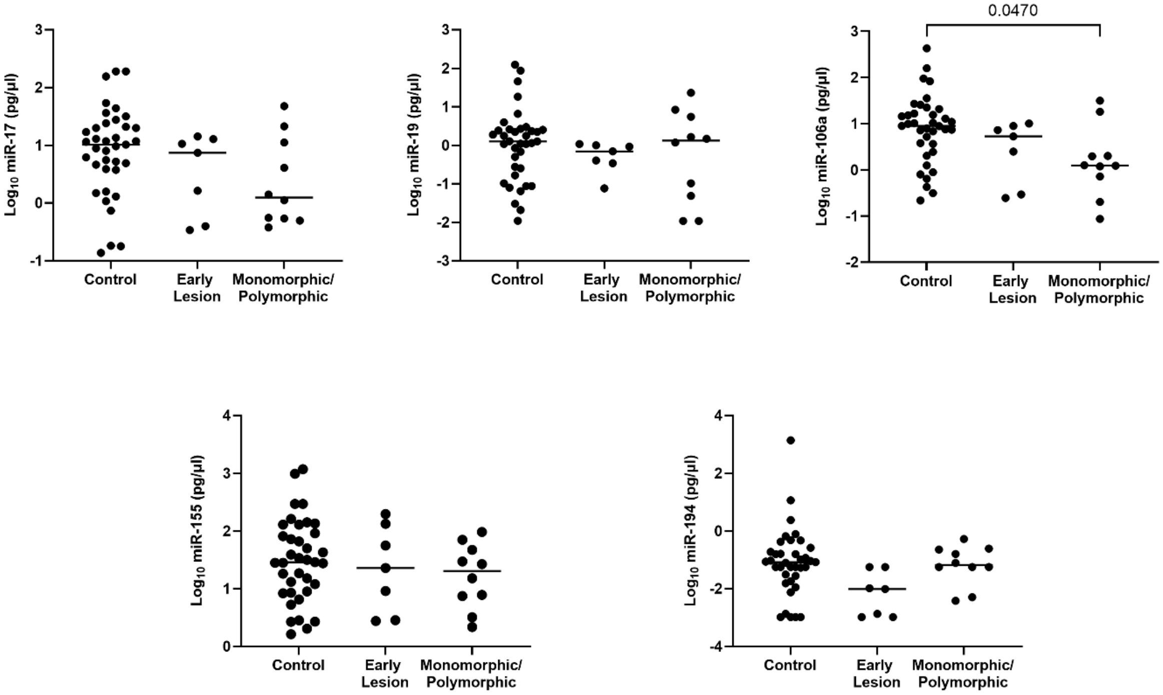


**Figure S1: Decreased levels of miRs are detected in the plasma of PTLD+ pediatric transplant recipients with monomorphic or polymorphic lesions as compared to controls.** Concentrations of miR-17, miR-19, miR-106a, miR-155 and miR-194 were measured by qPCR in plasma of PTLD+ patients with early lesions, monomorphic/polymorphic lesions and matched controls. miR-106a, controls vs monomorphic/polymorphic lesions, p = 0.047 by one-way ANOVA.

**Figure S2:**

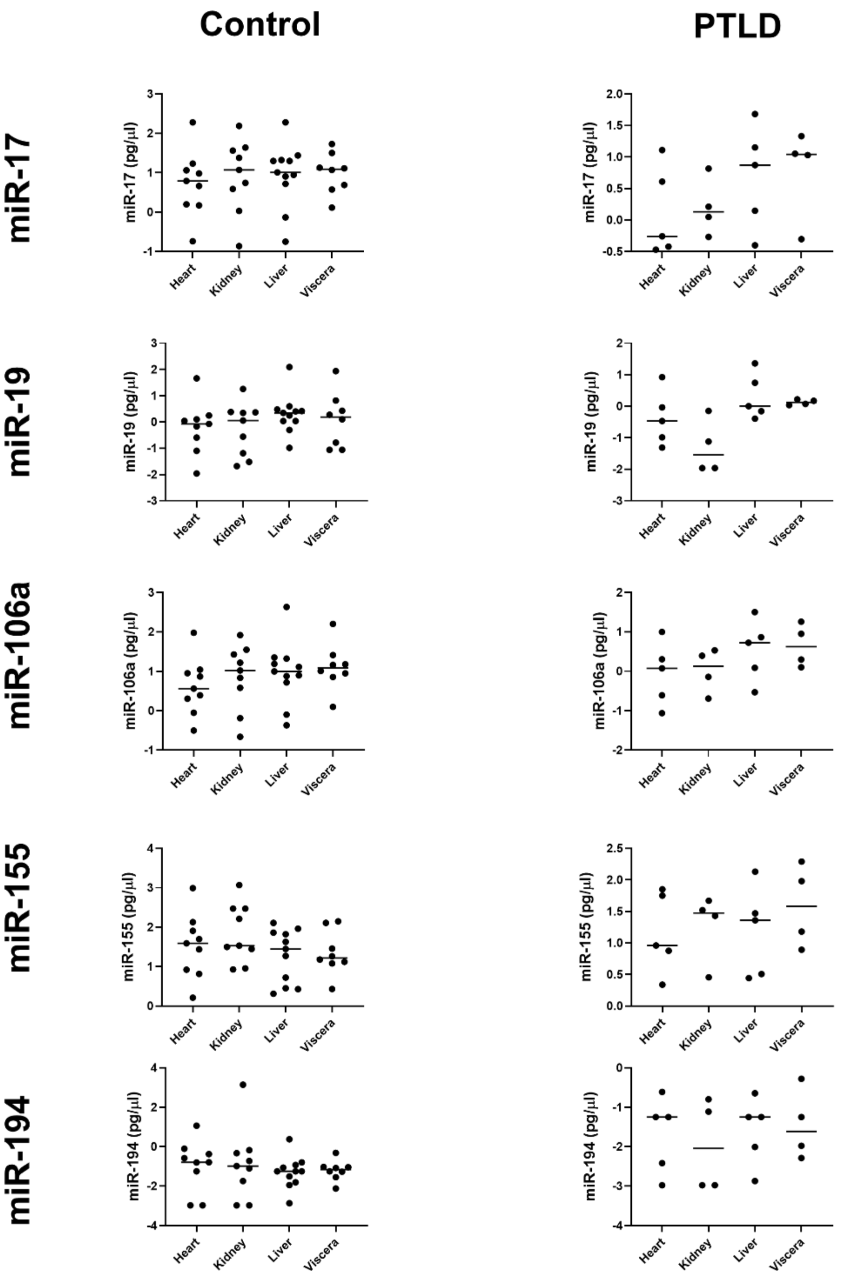


**Figure S2: The expression of miRs 17, 19, 106a, 155 and 194 did not differ significantly between the recipients of heart, kidney, liver and visceral transplants within PTLD+ and control groups.** Concentrations of miR-17, miR-19, miR-106a, miR-155 and miR-194 were measured by qPCR in plasma of recipients of heart, kidney, liver and visceral transplant.

### Figure S3:

**A.**

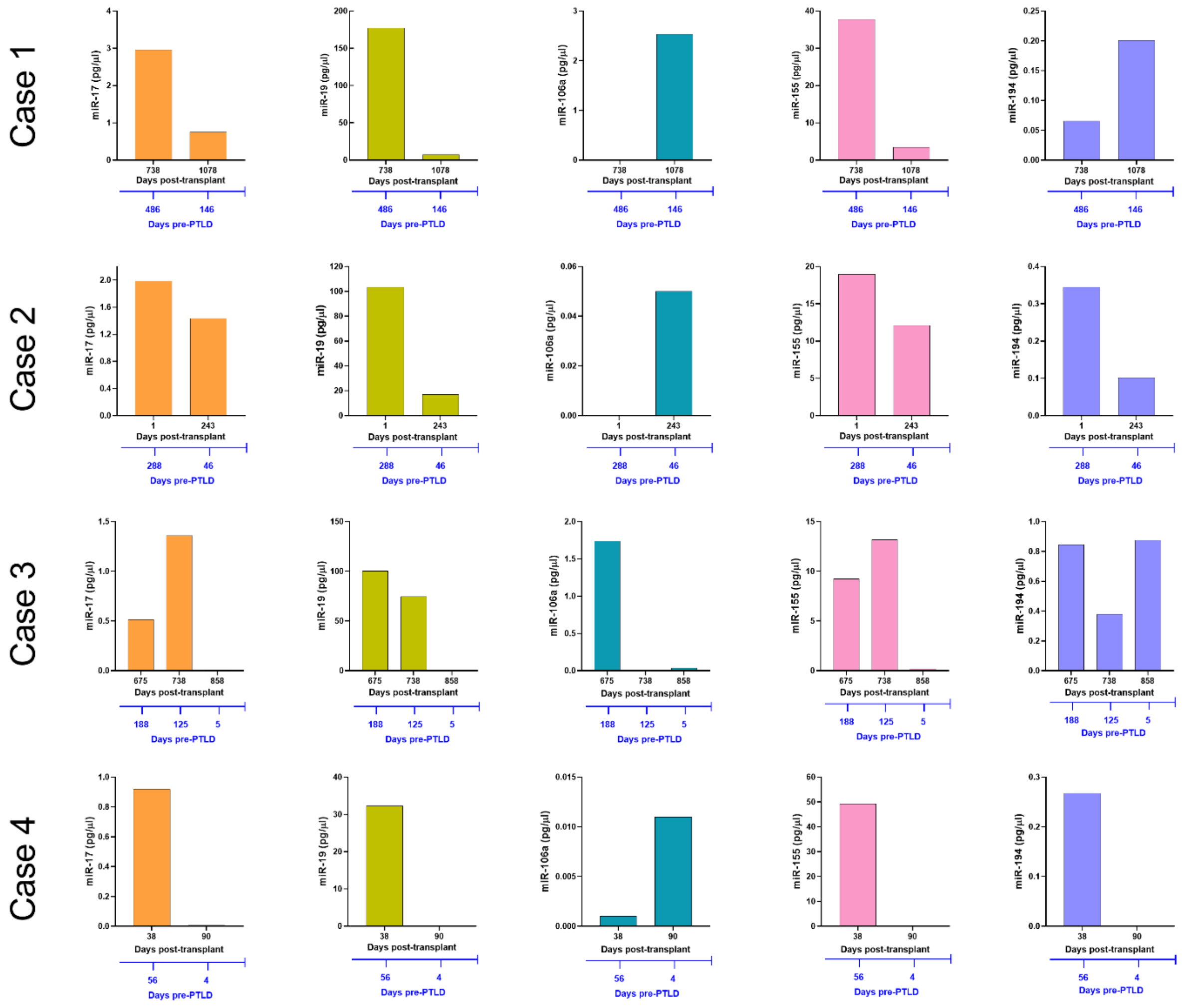
Case 1
Case 2
Case 3
Case 4
miR-17 (pg/µl)
miR-19 (pg/µl)
miR-106a (pg/µl)
miR-155 (pg/µl)
miR-194 (pg/µl)
Days post-transplant
Days pre-PTLD

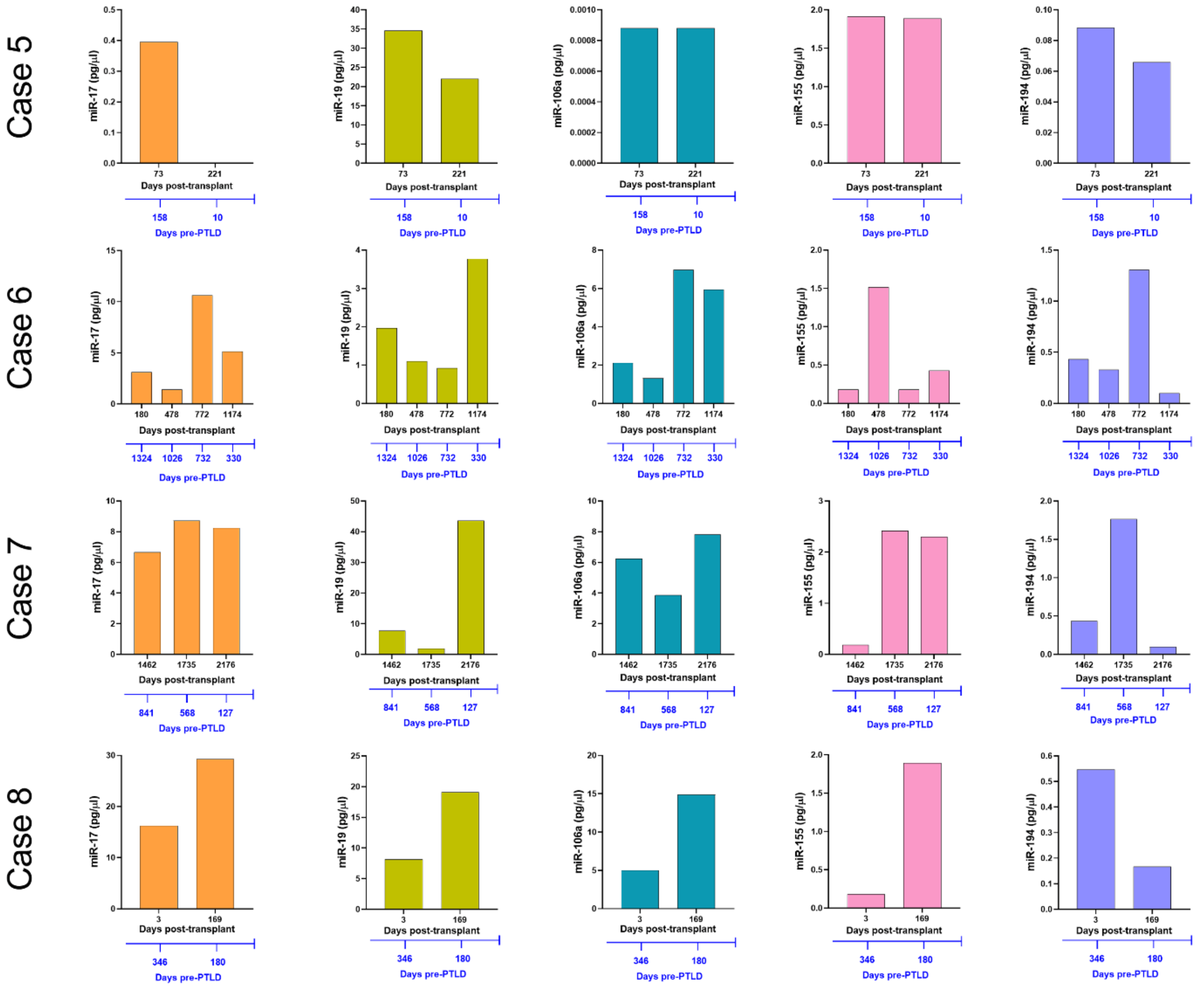
Case 5
Case 6
Case 7
Case 8
miR-17 (pg/µl)
miR-19 (pg/µl)
miR-106a (pg/µl)
miR-155 (pg/µl)
miR-194 (pg/µl)
Days post-transplant
Days pre-PTLD
73
221
158
10
180
478
772
1174
1324
1026
732
330
1462
1735
2176
841
568
127
3
169
346
180

**B.**

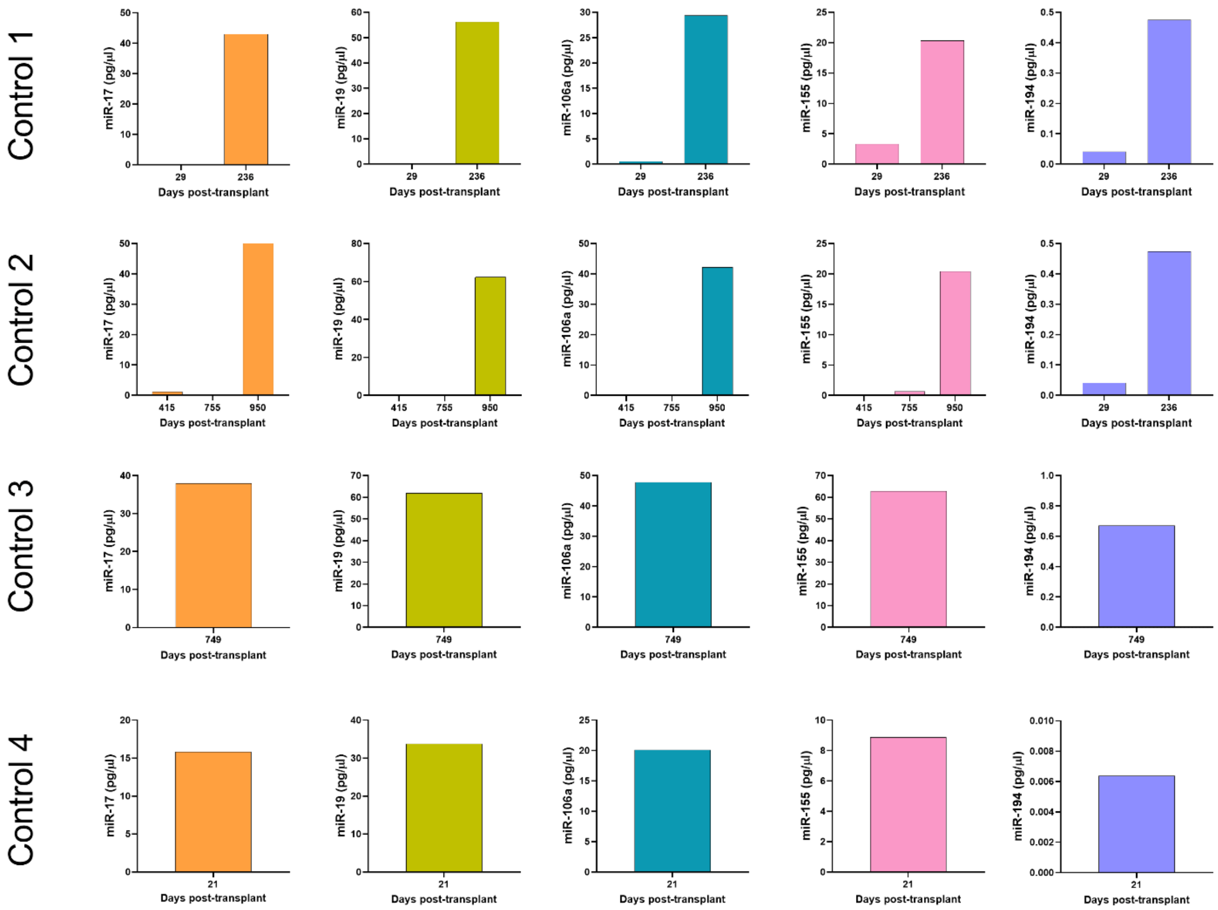

Control 1
miR-17 (pg/µl)
miR-19 (pg/µl)
miR-106a (pg/µl)
miR-155 (pg/µl)
miR-194 (pg/µl)
29
236
Days post-transplant
Control 2
miR-17 (pg/µl)
miR-19 (pg/µl)
miR-106a (pg/µl)
miR-155 (pg/µl)
miR-194 (pg/µl)
415
755
950
29
236
Days post-transplant
Control 3
miR-17 (pg/µl)
miR-19 (pg/µl)
miR-106a (pg/µl)
miR-155 (pg/µl)
miR-194 (pg/µl)
749
Days post-transplant
Control 4
miR-17 (pg/µl)
miR-19 (pg/µl)
miR-106a (pg/µl)
miR-155 (pg/µl)
miR-194 (pg/µl)
21
Days post-transplant

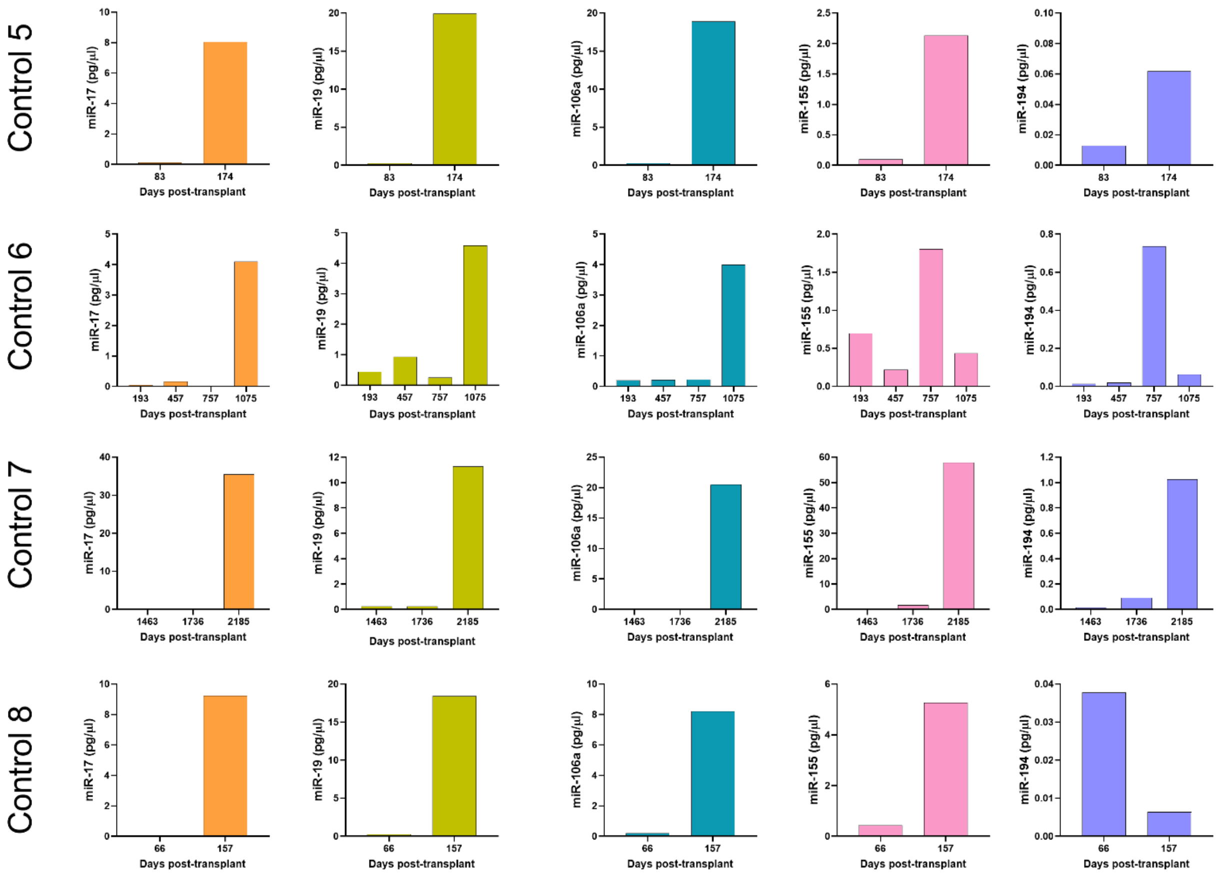


**Figure S3: Longitudinal analysis of plasma miRNA expression. (A)** Concentrations of miR-17, miR-19, miR-106a, miR-155 and miR-194 were measured by qPCR in plasma samples collected at multiple timepoints post-transplant from PTLD+ patients (n = 8). **(B)** Concentrations of miR-17, miR-19, miR-106a, miR-155 and miR-194 were measured by qPCR in plasma samples collected post-transplant from control patients at timepoints matched with paired PTLD+ patients (n = 8).

**<u>Figure S4:</u>**

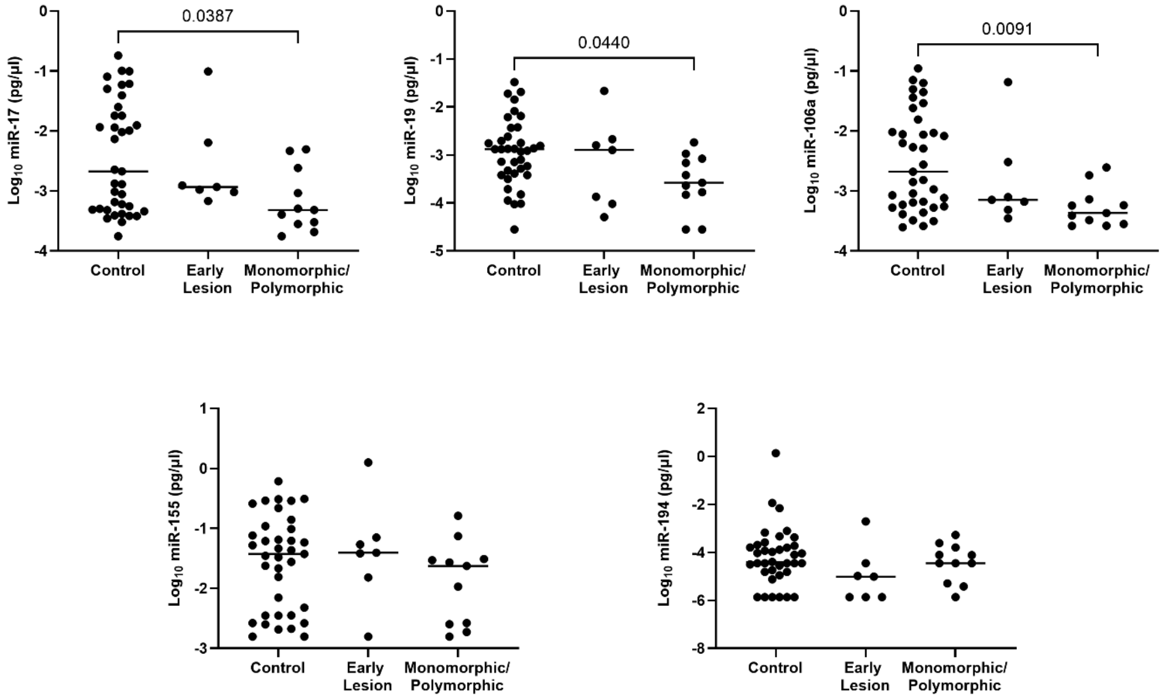


**Figure S4: Decreased levels of miRs are detected in the EV of PTLD+ pediatric transplant recipients with monomorphic or polymorphic lesions as compared to controls.** Concentrations of miR-17, miR-19, miR-106a, miR-155 and miR-194 were measured by qPCR in plasma of PTLD+ patients with early lesions, monomorphic/polymorphic lesions and matched controls. miR-17, controls vs monomorphic/polymorphic lesions, p = 0.039; miR-19, controls vs monomorphic/polymorphic lesions, p = 0.044; miR-106a, controls vs monomorphic/polymorphic lesions, p = 0.009, by one-way ANOVA.
